# The intra-household gendered burden of animal diseases in livestock-producing households in Ethiopia: Results from key informant interviews and a scoping review

**DOI:** 10.64898/2025.12.11.25341514

**Authors:** Eleanor Balchin, Theodore Knight-Jones, Luke O’Neill, Peregrine Rothman Ostrow, Sara Babo Martins, Wudu Temesgen, Jonathan Rushton

## Abstract

Ethiopia is a highly agrarian economy, though livestock’s contribution falls below its potential. Women play a significant role in livestock production; however, the literature on gendered dynamics of livestock disease is limited, particularly at the intra-household level. This work marks the first gender-focused study within the Global Burden of Animal Disease programme. Its goal is to enhance the programme’s aim of disaggregating the economic burden borne by humans due to animal disease. It explores the extent to which existing knowledge can be disaggregated by gender within households.

A scoping review of the existing literature on the intra-household burden of animal disease in Ethiopia was conducted, with 143 articles screened. This was supplemented by seventeen key informant interviews consisting of individuals or knowledgeable representatives from organisations known to the authors for their work in Ethiopia and/or on gender, livestock production, and animal disease.

Only one study directly addressed the intra-household gendered dimensions of animal disease burden in Ethiopia. Data were extracted in MS Excel. Adult men and women were found to be most impacted due to their roles in income generation and providing animal-sourced foods, and their need to compensate for losses during disease outbreaks. However, all household members contribute to disease transmission through gender-specific responsibilities. Key informant interviews were analysed in NVivo to determine themes in responses. Participants noted that household members engage in distinct transmission activities and face unequal consequences shaped by gendered norms and emphasised the need for gender-disaggregated data.

We advocate for primary, contextually grounded data collection that routinely includes gender- and age-disaggregated measures of exposure, decision-making, empowerment, and economic outcomes, complemented by qualitative enquiry. This would enable the design of targeted interventions to reduce animal and human morbidity and mortality while protecting livelihoods and promoting equity.

## Introduction

Ethiopia is a highly agrarian economy, with agriculture accounting for around 40% of GDP, 80% of exports, and employing approximately 75% of the country’s workforce [1]. Despite having the largest livestock population in Africa [2] livestock contributes less than 20% of the GDP, far below its potential [3]. Livestock also play a pivotal role at an intra-household level. They are a source of food, income, and employment; serve as a store of wealth; supply draught power and organic fertiliser for crop production, function as transport; and confer social status [4]. Animal disease limits this potential and can lead to losses and added expenditure [5].

Women represent two-thirds of poor livestock keepers worldwide [6] and are central to livestock production, yet robust analysis of gender within value chains is limited [7]. Existing literature is often low-quality regarding gender analysis [8], and lack depth in their consideration of topics such as ownership and local understandings of resource control [9].

Research specifically linking gender with the burden of livestock disease among livestock owners is sparse, and evidence at the intra-household level is rarer still. Understanding livestock disease from an intra-household perspective is essential to avoid generalisation across household members with different roles, exposures, and outcomes.

Available literature suggests that culturally patterned divisions of labour shape who is exposed to livestock diseases and how impacts are experienced, with roles varying by age [10–13]. These divisions influence which household member is positioned to notice disease symptoms and take preliminary action [10].

However, women’s ability to effectively manage animal disease may be limited. For instance, they lack decision-making power in important and high-value livestock transactions, including decisions related to the vaccination of livestock. They also experience reduced access to education and extension services, and restricted mobility due to social norms that influence where they can go, whom they can see, and how they can travel, limiting service access [14–16].

The consequences of animal disease are likewise unequally distributed within a household. Women spend around 90% of their incomes from agricultural production on their families, whilst men spend only 30-40% [17]. Therefore, a loss of livestock income controlled by women may affect children more greatly than a loss of men’s income. Women’s unpaid labour and care work may increase during animal health emergencies, and their assets may be the first to be sold in response to such events [18].

Much of the current evidence, monitoring, and burden estimation still operate at the household level, and utilise a monetary-focused framework for burden estimation [19]. This fails to capture the full spectrum of impacts and often neglects the intra-household differences in exposure and burden, which obscures gender and power dynamics. The importance of gender differentiation in this context has led the Global Burden of Animal Disease (GBADs) programme to seek ways to disaggregate animal disease burdens by gender of household members. Building on the Ethiopian case study work within GBADs, this review addresses key evidence gaps identified in previous research globally. The extent of this gap lends this study to a scoping review and key informant interviews. The scoping review element allows us to identify the extent and nature of existing evidence, whilst key informant interviews bring experiential and tacit knowledge from practitioners, policy makers or researchers who understand the realities of the field of study.

The review highlights the persistent inequities and priorities for gender-responsive interventions in Ethiopian livestock systems. It aims to inform gender-responsive policies and interventions for livestock health in Ethiopia and similar settings.

### Objectives

The objective of this study was to systematically review and synthesise current knowledge regarding the intra-household, gender-differentiated burden of animal diseases in Ethiopia through a scoping review and key informant interviews (KIIs). Specifically, the study aimed to document existing research specifically documenting how gender affects exposure to livestock disease, gendered roles in the transmission of livestock disease, and how livestock disease affects household members in different ways.

## Methodology

Employing a mixed-method approach, this study was informed by a pragmatic research paradigm. A scoping review and KIIs were integrated to allow us to explore the breadth and depth of existing knowledge on this under-researched subject and determine what data available for disaggregation at the intra-household level.

### Scoping review

The review protocol was guided by the work of Arksey and O’Malley [20]. The electronic databases Scopus, Web of Science, and PubMed were searched for relevant studies. The search strings were adapted using keywords relating to intra-household, gender, disease, impact, livestock and Ethiopia for use in each database, available in the OSF Registries at https://doi.org/10.17605/OSF.IO/K5JEC. Field tags for title and abstract were applied, requiring two separate searches in Web of Science.

An example of the full electronic search strategy for one of the databases, Scopus, is as follows:

TITLE-ABS (gender OR women OR woman OR “household dynamics” OR “intrahousehold” OR “intra-household” OR “intra household”) AND TITLE-ABS (illness OR disease OR burden OR impact OR transmission OR “food safety” OR infection OR zoonosis OR zoonoses OR “food borne disease” OR “infectious disease” OR antimicrobial) AND TITLE-ABS (livestock OR chicken OR turkey OR geese OR poultry OR sheep OR goat OR “small ruminant” OR pig OR cattle OR cow OR bovine OR heifer OR bull OR calf OR calves OR steer OR ox OR camel OR equid OR horse OR donkey OR ruminant) AND TITLE-ABS (Ethiopia)

All document types and only results in English were included to allow for the broadest range of results possible, acknowledging the language abilities of the authors. A cutoff date of 31/12/2023 was applied, to allow the authors time to analyse and report the results.

To obtain relevant papers, inclusion and exclusion criteria were employed during each eligibility phase (Table 1).

**Table 1:** Inclusion and Exclusion Criteria.

| **Title Review** |  |
| --- | --- |
| Livestock production systems (including management and productivity), gender roles in agricultural production. | No connection to livestock or their related diseases |
| Livestock's role in food security and nutrition | Purely veterinary or epidemiology or laboratory studies with no link to households or humans (e.g. seroprevalence studies, vaccine development, etc.) |
| Animal welfare | Relating to other countries |
| Feed safety and animal disease transmission |  |
| Safety nets (e.g. social protection, resilience programmes) in relation to agriculture |  |
| **Abstract Review** |  |
| Impact of livestock disease on household members | No connection to livestock disease, or impact on households |
| Risk of disease transmission in households | Relating to other countries |
| Household coping strategies in response to disease |  |
| Gendered disease prioritisation |  |
| **Full-Text Review** |  |
| Explicitly disaggregates at the intra-household level disease exposure or impacts | Insufficient depth to disaggregate at the intra-household level |

The first author (EB) led the process of performing the search, developing the criteria, and charting the results. A second reviewer (LON) then confirmed the findings by independently screening at random 10% of the results at each screening stage.

Only full-text studies written in English were included. Data were extracted and analysed using Microsoft Excel to determine the topics discussed in the literature. Studies were included if they contained gender-disaggregated data at the intra-household level relating to exposure to or impacts of animal disease. The scoping review portion of this study is reported following the PRISMA Extension for Scoping Reviews (PRISMA-ScR) checklist [21].

### Key informant interviews

KIIs were conducted between the 8^th^ February and 13th March 2023, under ethical approval reference ILRI-IREC2022-14/1. Interviews were conducted either in person at the International Livestock Research Institute (ILRI) in Addis Ababa, at the institution of the key informant (KI), or through video conferencing, whichever was most convenient for the participant, to foster a relaxed discussion around the research questions and acknowledge logistical realities where KIs are based in other countries. We used purposive sampling to recruit seventeen KIs based on the authors’ knowledge of their work in Ethiopia and/or on gender, livestock production, and animal disease. They were perceived as having ‘special or expert knowledge’ [22] in these areas, and were drawn from a range of relevant sectors, including government, academia, NGOs and international organisations and included both male and female respondents. Participant recruitment occurred on a rolling basis between 5^th^ January to 1^st^ March 2025. Participants completed a written consent form before their interviews which included an overview of the study and how the interview data will be used, information on how and where data will be stored, and had the option to agree or not agree to audio recording.

The KIs were asked 3 main questions:

1. Is there a difference in how household members are exposed to animal diseases?
2. Is there a difference in how household members are impacted by disease?
3. What are the gaps in either the literature or interventions relating to the gendered burden of animal diseases?

EB and TK-J conducted the KIIs, with EB present at all interviews, and TK-J supporting earlier interviews. Sixteen of the seventeen KIs consented to audio recording. Brief notes were taken during the interviews and were expanded on the same day whilst listening to the recording. This allowed the interviewer to focus more on the discussion than on note-taking [23]. For the interview where consent for audio recording was not granted, more extensive notes were taken immediately following the interview [24]. Audio recordings were stored on the University of Liverpool’s secure server and were deleted once the contracted project period ended in May 2023. Data were coded by EB using NVivo to establish the themes that were discussed and categorise responses. The key informant portion of this study is reported here in accordance with the Standards for Reporting Qualitative Research (SRQR) checklist [25].

## Results

### Scoping review

Of 143 unique studies identified, only one by Gizaw et al (2020) [26] collected data related to the gendered burden of disease within households in Ethiopia. Fig 1 shows the scoping flow diagram through the review process.

**Fig 1:**
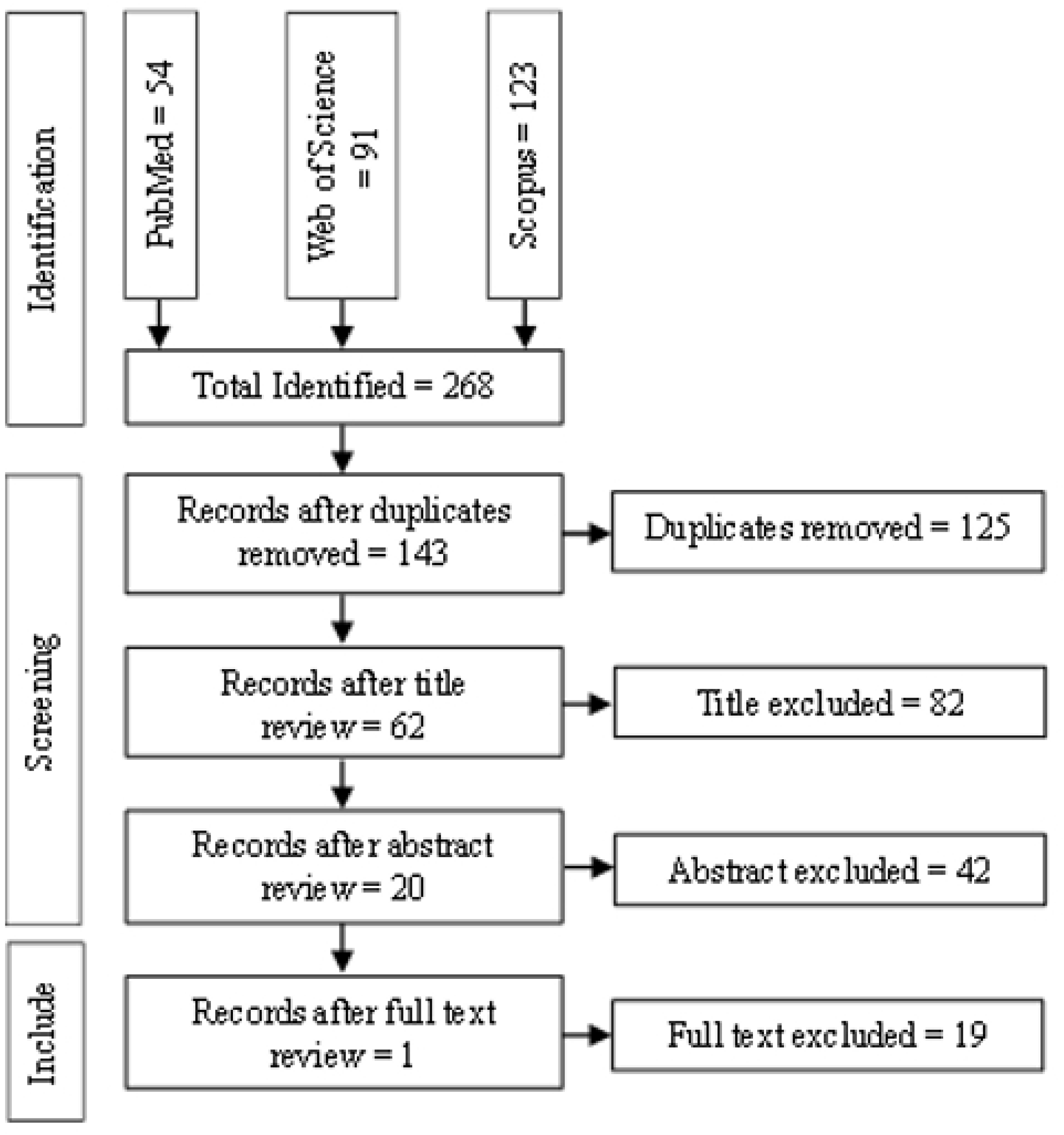
Scoping Review Diagram

The included paper was conducted in Amhara, Oromia, Southern Nations and Nationalities and Peoples region (SNNRP), and Tigray, across 17 districts and 37 kebeles (the smallest administrative units). Seventy focus group discussions (FGDs) were conducted, involving separate male and female groups in each of the kebeles. Two kebeles did not have male focus groups, three didn’t have female focus groups, and one had two female focus groups. FGDs involved group discussion and used proportional piling to elicit farmer priorities and perceptions.

Data from proportional piling results showed the intra-household roles in livestock production that may be linked to disease transmission and illustrated how certain household members may be affected by disease. However, these results are not comprehensively examined or discussed. Instead, analysis and discussion centred more around male versus female disease priority data that were collected.

### Key informant interviews

#### Gendered disease transmission risk

Table 2 summarises the key informants’ understanding of the association between each household member and animal disease transmission. It details the animals thought to be the responsibility of each member or those they are able derive benefits from; roles in animal production that may contribute to disease transmission; consumption-related risks specific to each household member; and diseases most likely to affect each member of the household, either through zoonotic transmission to humans, or as a result disease burden affecting the livestock they manage or benefit from. The KIs were not asked about differences in transmission and impact of zoonotic and non-zoonotic diseases.

**Table 2:** Overview of Key Informants Understanding of Intra-Household Gendered Burden of Disease.

|  | Women | Men | Girls | Boys |
| --- | --- | --- | --- | --- |
| **Animal species associated with the household member** | • Chicken<br>• Small Ruminants<br>• Dairy Cattle<br>• Donkeys | • Cattle<br>• Oxen<br>• Camels | • Chicken<br>• Small Ruminants<br>• Dairy Cattle<br>• Donkeys | • Chicken<br>• Small Ruminants<br>• Dairy Cattle<br>• Donkeys<br>• Cattle<br>• Oxen |
| **Production activities that pose disease risks** | • Cleaning Shelters<br>• Feeding Livestock<br>• Caring for Sick Animals | • Herding<br>• Marketing<br>• Slaughtering<br>• Removing hides<br>• Disposing of sick animals | • Often follow the mother's role | • Often follow the father's role, as well as assisting mother |
| **Consumption of animal-sourced foods particularly affecting disease exposure** | • Consume less animal-sourced food than men and children | • Raw Meat | • Raw Milk | • Raw Milk |
| **Diseases to which household member is most vulnerable** | • Avian Influenza<br>• Campylobacter<br>• Brucellosis<br>• Hydatidosis<br>• Rabies<br>• Tuberculosis<br>• Toxoplasmosis | • Anthrax<br>• Brucellosis<br>• Blackleg | • Avian Influenza<br>• Brucellosis<br>• Salmonellosis | • Anthrax<br>• Brucellosis<br>• Blackleg |

Transmission activities associated with women included milking, cleaning shelters, caring for sick livestock, slaughtering chickens, preparing raw meat for cooking, and assisting in births, particularly in relation to poultry, small ruminants, dairy cattle, and donkeys. When employed at slaughterhouses, women were seen to be involved in cleaning, which often lacks sanitisation, involving simply rinsing with water. Women were said to spend most of their time in their households, resulting in close contact with animals.

Women were thought to eat less animal-source food than other household members. When they do eat animal-sourced foods it is thought that they tend to eat different parts of the animal (as one participant explained, meat is for men, offal is for women). KI described how in some areas in Ethiopia, taboos around women eating certain foods, such as camel milk or eggs, result in geographically differing exposure to zoonotic disease through consumption.

Women were thought to be particularly at risk of exposure to and/or the impacts of, avian influenza, brucellosis, campylobacter, hookworm, hydatidosis, rabies, salmonellosis, tuberculosis, and toxoplasmosis. These are zoonotic diseases present in poultry, small ruminants, dairy cattle, cats and dogs, which are more within women’s sphere. Women are thought to be responsible for the care of, be in the closest contact with, and/or able to control the income from these animals.

Identified transmission activities among men included herding, marketing, slaughtering, and consumption of more meat and blood products. Men are more often employed at slaughterhouses in roles other than cleaning and tend to slaughter the livestock at home. They are responsible for the larger livestock species and are involved in agricultural roles that take them away from the household. Men are responsible for removing the hide and disposing of dead livestock, and some KIs raised the issue of the consumption of meat from dead animals if the farmers believed it to have died from ‘a normal disease’. Men were also thought to be first in line for eating animal-sourced food and so may be more at risk of contracting disease this way.

The diseases that men were seen to be most at risk of exposure to and/or the impacts of were anthrax and brucellosis due to consumption habits and close contact with cattle, whilst blackleg was seen to result in significant economic losses for men.

Identified transmission activities among children included the consumption of milk, cleaning animal shelters, water collection, herding (for boys), general close contact with animals such as playing with dogs, cats, and chickens at the household and their regular proximity to the household.

Most often discussed by the KIs was children’s relatively high consumption of raw milk, with mention of how children drink milk directly from an animal’s udders at times. Children’s roles in livestock production were seen to put them at risk of spreading disease across animals and exposure to zoonotic disease through direct contact. One KI also raised how in some culture’s boys drink raw blood at their initiation ceremonies.

Children were considered to be most at risk of exposure to and/or the impacts of rabies, hydatidosis, toxoplasmosis, brucellosis, and avian influenza. In addition, girls were thought to be at risk of exposure to and/or the impacts of salmonellosis due to their roles caring for animals, particularly chickens. Boys, on the other hand, were thought to be at risk of exposure to and/or the impacts anthrax and blackleg due to their roles in managing cattle.

#### Gendered impacts of animal disease

The KIs agreed that different members of a household are impacted by different diseases in different ways. Table 3 shows the ranking of the number of informants discussing a certain topic. The KIs identified the most extensive variety of impacts of animal disease in relation to women.

**Table 3:** Gendered Impacts of Animal Disease Within the Household as Identified by the KI’s.

| Rank | Women | Men | Children General | Girls | Boys |
| --- | --- | --- | --- | --- | --- |
| 1 | Increased labour burden | Income loss | School absenteeism | School absenteeism | No specific impacts raised |
| 2 | Income loss | Increased labour burden | Loss of nutrition | Increased labour burden |  |
| 3 | Loss of nutrition |  |  |  |  |
| 4 | Impact on wider family |  |  |  |  |
| 5 | Loss of empowerment |  |  |  |  |
| 6 | Psychological impact |  |  |  |  |
| 7 | Loss of well-being |  |  |  |  |
| 8 | Positive impacts of reduced number of animals |  |  |  |  |

Most often, women’s increased labour burden was seen to result from morbidity and mortality in donkeys, and their responsibility to care for sick animals. Donkeys were considered to be key in women’s domestic work, assisting in water collection and transporting goods to market. Morbidity that results in the donkey being unable to work, and mortality, were considered to result in women having to fulfil the donkey’s roles themselves, with several KIs stating that “*when a donkey is lost, a woman becomes the donkey”*. Women’s labour burden was also seen to increase as a result of having to care for sick livestock and household members, while covering additional daily duties when necessary.

The loss of income that results from disease in dairy cattle, small ruminants, and poultry was thought to significantly impact women. These livestock were thought to be those from which women have the potential to gain and control income. One KI spoke of how women’s incomes may be less diversified than men’s, and so loss of yields or death of livestock that results in reduced income can be more significant and more difficult to replace for women than for men. Women’s loss of income was also linked to their financial responsibilities within the household, such as paying for children’s clothing, school needs, and food.

Loss of nutrition was seen to impact women more than men or children, as women were seen to be last in line for food in the household. One KI stated the hierarchy for food provision within a household is men-children-women and even pregnant women were said to be last in line for food.

Women being impacted by animal disease themselves was said to have a significant impact on the wider family due to their vital role in domestic and farm work in two ways: wider impacts that result from women’s inability to conduct their agricultural and household duties, and impacts from women’s loss of income due to morbidity or mortality of the animals from which they are able to control income. Women’s illness was thought to impact children, and daughters in particular, as they were expected to take on their mother’s duties. Women’s loss of income, particularly livestock-related income, was thought to have a significant negative effect on children, and again, daughters in particular, as women spent the vast majority of their income on nutrition, education, and healthcare.

The loss of empowerment discussed by some KIs was related to the loss of decision-making power and control women have within the household. Women were thought to have more power over dairy cattle, small ruminants, and poultry, and it was said that if these livestock die, women may lose their control over income, and power within the home.

Some KIs believed that women suffer a psychological impact and loss of well-being as a result of their loss of livestock and status, increased workload, and the stress that comes with this, along with their loss of decision-making power within the household. A reduction in women’s life span as a result of their efforts at home was raised by one KI.

One notable insight highlighted the potential positive impact of reduced animals due to mortality, related to the considerable burden often borne by women. In reference to the potential reluctance of women to vaccinate their livestock, a KI explained how not vaccinating cattle lessened the growth of the herd, and thus prevented an increase in women’s labour burden, and reduced the risk of men taking over dairy production and keeping profits for themselves.

Fewer topics were raised in relation to how men are impacted by animal disease. The KIs only considered men to be impacted in terms of income loss and increased labour burden. Most often the loss of income was seen to be due to men controlling incomes from large livestock such as cattle and oxen, as these livestock command a high price. Additionally, loss of draught power was seen to impact only men, and loss of working equids for market transport seen to impact both men and women. Men’s increased labour burden was also seen to be due to the loss of draught power from oxen.

No KI provided any impacts that specifically affect boys. Most often, children were considered to be impacted by school absenteeism as a result of the increased labour burden of children assisting with tasks such as water collection, and loss of income. Loss of nutrition, particularly due to reduced milk yields, was thought to be a significant impact to children due to the role of animal source food consumption improving nutrition and potentially reducing stunting. School absenteeism was seen to be a particular concern for girls, as they are more likely to be removed from school to care for livestock and the home even in the absence of disease.

Girls are thought to often be involved in the same roles in the household as their mothers, and so when a woman is impacted by increased labour burden or when the mother becomes unwell, girls labour burden also increases, and so they are removed from school.

#### The need for further research

The majority of KIs believed that considerably more research is needed to understand the intra-household burden of animal disease. Half the KIs stated that more research is required to understand gender dynamics within a household, with gender roles and decision-making power being discussed most frequently. Many believed there is a severe lack of gender disaggregated data in animal health and infectious disease research. It was thought a deeper understanding of gender empowerment and equality, why intra-household data collection is important, and how it should be collected were key aspects of future work that should be disseminated among researchers. This knowledge is also required to allow gender-specific interventions to compete for resources and to inform government policy.

In order to provide strong data, many KI’s believed that GBADs must focus on primary data collection. The small number of studies that exist relating not only to the gendered burden of animal disease, but also to gender and livestock production more generally, limits GBAD’s ability to estimate the disaggregated burden of animal disease. The lack of research on this subject results in an inability to further disaggregate data by geography, culture, farming systems, disease, and animal species. Furthermore, some existing gendered data were considered weak and of low quality. Overall, these sentiments collectively led to a very low confidence should GBADs attempt to disaggregate gendered burden of animal disease from this existing data.

In addition, KIs often stated that quantitative economic data alone is not enough to be able to understand the gendered burden of animal disease. Rather, qualitative studies of the social sciences must be utilised in order to provide a full understanding of how household members are burdened and what secondary impacts this has. In all, there was concern that this masking of realities on the ground could lead to misinformed, unsuccessful, and inappropriate interventions. In line with this, GBADs ability to recommend methodologies for the collection of this primary data was considered one way in which the programme could be particularly valuable.

#### Considerations for improvement to animal health services and training

In addition to improving available data, the KI’s also raised that interventions need improvement in relation to animal health services and training. Trainings available to communities were considered to be both lacking, and not inclusive. Women’s domain was seen to be within the household, with high labour burdens limiting leisure time. In some communities where the cultural norm prevents women from travelling far from their households, women may be unable to attend available trainings, and men may not pass information learnt during trainings on to women.

It was noted that nationally, men tend to be the providers of animal health services due to their generally higher levels of education, which may reduce women’s use of these services due to cultural norms restricting women’s interactions with men or feelings of discomfort.

## Discussion

### Understanding intra-household impacts of animal disease

Understanding the impact of animal disease on household members requires consideration of how individuals interact with their livestock, the role of these animals in everyday life, and how household members manage shocks. This review has highlighted key themes in these relationships, particularly the importance of intra-household dynamics in shaping how disease is experienced. Research on gender and livestock production is scarce, comparing men and women more generally or household headship where it does exist [27]. There is an even greater lack of research considering the gendered burden of animal disease within households in Ethiopia, with only one existing study identified.

Drawing on both the proportional piling results from Gizaw et al (2020) and the KI insights, we suggest that the impacts of animal disease be grouped into three categories: Health and well-being, economic, and livelihood impacts. The extent to which each household member is exposed to animal disease and affected by economic or social consequences of disease reflects not only their labour roles, but also household power dynamics, access to resources, and decision-making authority, all of which are shaped by gender, age and cultural context.

Table 4 provides a visual summary of the results found in this review and how gender and intrahousehold dynamics relate to animal disease more broadly. Specifically, it illustrates which animals were identified as being within the realm of each household member, disease transmission activities associated with each household member, and the direct and indirect impacts identified as being of particular concern for each. The results suggest that there are direct economic health and well-being-related impacts of animal disease; namely a loss of income (primarily experienced by men and women), gendered exposure to zoonoses (affecting all household members), as well as loss of nutrition and food security (affecting primarily women and children) as a direct result of yield reductions of animal-sourced foods. Indirect impacts related to health and well-being, and livelihoods, including increased labour burdens, psychological impacts, and decreased school attendance. Health and well-being impacts included a detraction of psychological and emotional well-being which were thought to particularly affect women, whilst indirect loss of nutrition and food insecurity resulting from loss of income was thought to further jeopardise the health of women and children. Finally, indirect impacts to people’s wider livelihoods included children’s school absenteeism, a reduction of women’s empowerment, and increased or altered labour burdens, particularly affecting women, men, and young girls.

**Table 4:**
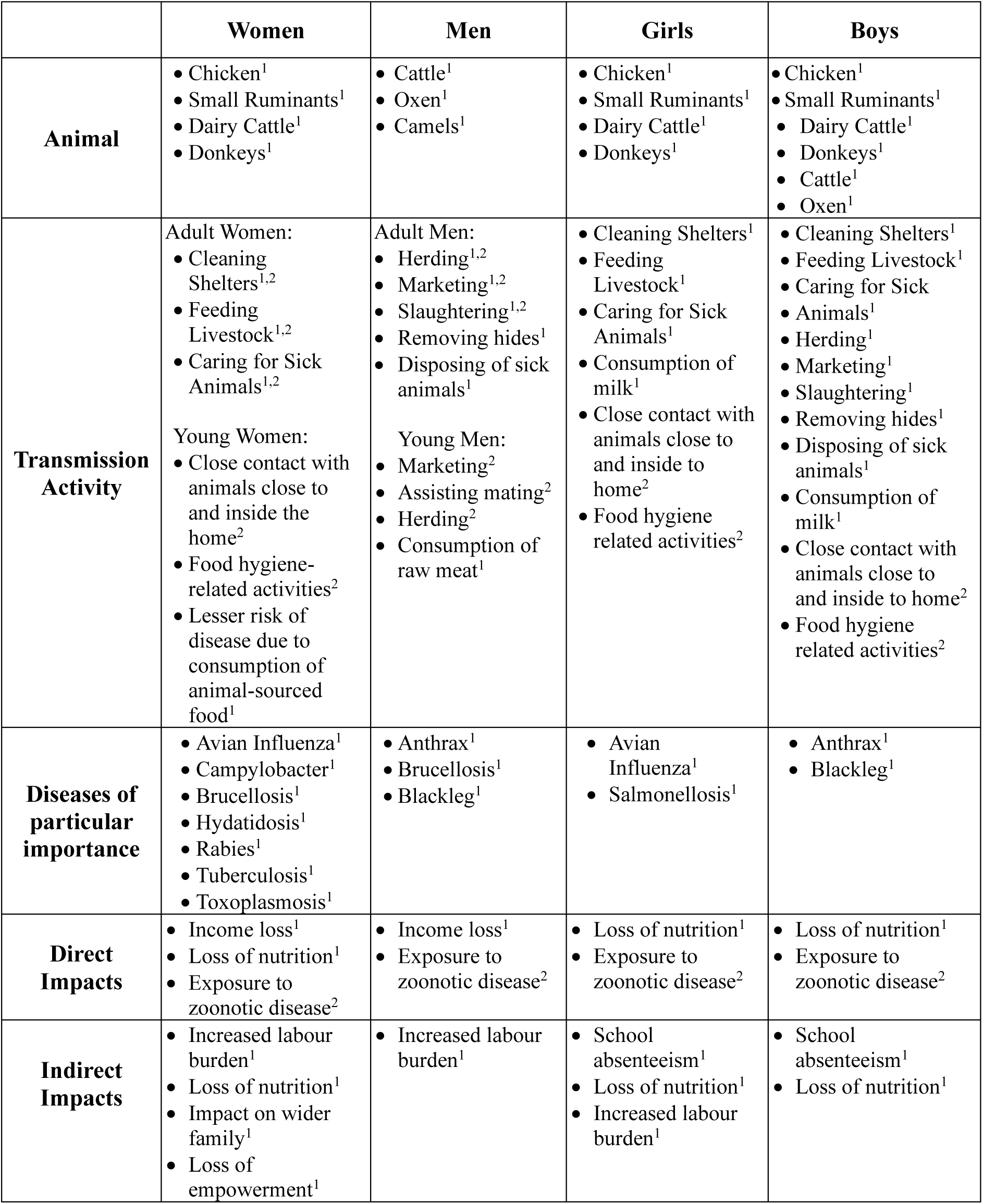
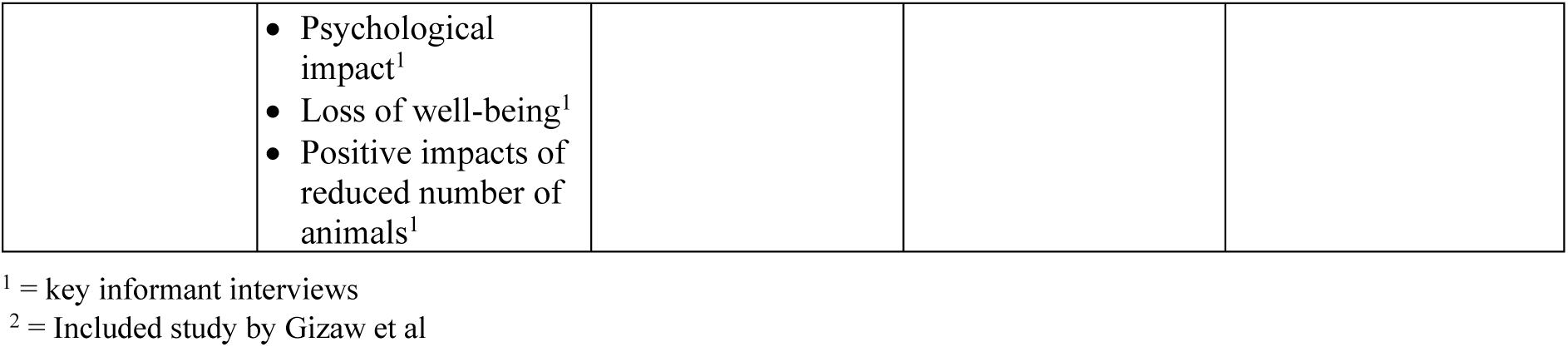
Combined Results from KIs and Scoping Review Mapping What is Currently Known About the Intrahousehold Burden of Animal Disease in Ethiopia.

|  | **Women** | **Men** | **Girls** | **Boys** |
| --- | --- | --- | --- | --- |
| **Animal** | • Chicken<sup>1</sup><br>• Small Ruminants<sup>1</sup><br>• Dairy Cattle<sup>1</sup><br>• Donkeys<sup>1</sup> | • Cattle<sup>1</sup><br>• Oxen<sup>1</sup><br>• Camels<sup>1</sup> | • Chicken<sup>1</sup><br>• Small Ruminants<sup>1</sup><br>• Dairy Cattle<sup>1</sup><br>• Donkeys<sup>1</sup> | • Chicken<sup>1</sup><br>• Small Ruminants<sup>1</sup><br>• Dairy Cattle<sup>1</sup><br>• Donkeys<sup>1</sup><br>• Cattle<sup>1</sup><br>• Oxen<sup>1</sup> |
| **Transmission Activity** | Adult Women:<br>• Cleaning Shelters<sup>1,2</sup><br>• Feeding Livestock<sup>1,2</sup><br>• Caring for Sick Animals<sup>1,2</sup><br>Young Women:<br>• Close contact with animals close to and inside the home<sup>2</sup><br>• Food hygiene-related activities<sup>2</sup><br>• Lesser risk of disease due to consumption of animal-sourced food<sup>1</sup> | Adult Men:<br>• Herding<sup>1,2</sup><br>• Marketing<sup>1,2</sup><br>• Slaughtering<sup>1,2</sup><br>• Removing hides<sup>1</sup><br>• Disposing of sick animals<sup>1</sup><br>Young Men:<br>• Marketing<sup>2</sup><br>• Assisting mating<sup>2</sup><br>• Herding<sup>2</sup><br>• Consumption of raw meat<sup>1</sup> | • Cleaning Shelters<sup>1</sup><br>• Feeding Livestock<sup>1</sup><br>• Caring for Sick Animals<sup>1</sup><br>• Consumption of milk<sup>1</sup><br>• Close contact with animals close to and inside to home<sup>2</sup><br>• Food hygiene related activities<sup>2</sup> | • Cleaning Shelters<sup>1</sup><br>• Feeding Livestock<sup>1</sup><br>• Caring for Sick Animals<sup>1</sup><br>• Herding<sup>1</sup><br>• Marketing<sup>1</sup><br>• Slaughtering<sup>1</sup><br>• Removing hides<sup>1</sup><br>• Disposing of sick animals<sup>1</sup><br>• Consumption of milk<sup>1</sup><br>• Close contact with animals close to and inside to home<sup>2</sup><br>• Food hygiene related activities<sup>2</sup> |
| **Diseases of particular importance** | • Avian Influenza<sup>1</sup><br>• Campylobacter<sup>1</sup><br>• Brucellosis<sup>1</sup><br>• Hydatidosis<sup>1</sup><br>• Rabies<sup>1</sup><br>• Tuberculosis<sup>1</sup><br>• Toxoplasmosis<sup>1</sup> | • Anthrax<sup>1</sup><br>• Brucellosis<sup>1</sup><br>• Blackleg<sup>1</sup> | • Avian Influenza<sup>1</sup><br>• Salmonellosis<sup>1</sup> | • Anthrax<sup>1</sup><br>• Blackleg<sup>1</sup> |
| **Direct Impacts** | • Income loss<sup>1</sup><br>• Loss of nutrition<sup>1</sup><br>• Exposure to zoonotic disease<sup>2</sup> | • Income loss<sup>1</sup><br>• Exposure to zoonotic disease<sup>2</sup> | • Loss of nutrition<sup>1</sup><br>• Exposure to zoonotic disease<sup>2</sup> | • Loss of nutrition<sup>1</sup><br>• Exposure to zoonotic disease<sup>2</sup> |
| **Indirect Impacts** | • Increased labour burden<sup>1</sup><br>• Loss of nutrition<sup>1</sup><br>• Impact on wider family<sup>1</sup><br>• Loss of empowerment<sup>1</sup> | • Increased labour burden<sup>1</sup> | • School absenteeism<sup>1</sup><br>• Loss of nutrition<sup>1</sup><br>• Increased labour burden<sup>1</sup> | • School absenteeism<sup>1</sup><br>• Loss of nutrition<sup>1</sup> |
|  | • Psychological impact<sup>1</sup><br>• Loss of well-being<sup>1</sup><br>• Positive impacts of reduced number of animals<sup>1</sup> |  |  |  |

<sup>1</sup> = key informant interviews
<sup>2</sup> = Included study by Gizaw et al

Whilst results on transmission activity presented by Gizaw et al (2020) suggest defined divisions in exposure risk in the included study, some FGD groups reported that adult men were the most impacted by livestock disease, others reported adult women being more affected. Some saw the impact as equal between the two. This discrepancy may reflect the difference between perceived impact and actual exposure and burden, or men and women may be similarly affected overall. Still, the nature of that impact may diverge based on their roles in livestock production and within the household, as was suggested by the KIs. This divergence also demonstrates the value of using participatory tools such as proportional piling to reveal hidden or underestimated exposure pathways.

### Economic impacts of animal disease

#### Loss of income

Livestock diseases can have substantial economic implications for both men and women in livestock farming households, as evidenced in the results of both this scoping review and the KIIs. These households often rely heavily on the incomes generated from their livestock, making morbidity and mortality due to disease a critical concern, with cascading effects on the entire household. Livestock diseases directly impact livestock productivity by causing reduced birth rates and increased mortality, alongside decreased milk production, animal offtake, and draught efficiency.

Previous studies have sought to quantify the economic losses associated with animal disease at the household level [28–30]. However, a significant gap exists in the literature regarding the understanding of how these economic impacts are distributed within a household. Our results suggest that income losses affect men and women in different ways. While both are shown to be involved in income-generating activities, adult men tend to dominate market-facing activities, such as marketing and slaughter. In contrast, women are more involved in caregiving and feeding roles. Women may face greater challenges in compensating for lost income due to reduced opportunities for income generation, along with variations in financial responsibilities within the household. For example, women often allocate more of their income to essential needs like food [31,32], healthcare [33], and education [34], which influences how the burden of lost income due to disease is distributed within the household.

### Health and well-being impacts of animal disease

#### Gendered exposure to zoonotic disease

Gendered exposure to zoonotic disease is a central concern when assessing the intrahousehold burden of animal disease. Gendered roles in livestock production, which vary across production systems, are well-documented [8,27,35–37], and some studies have explored how these roles influence an individual’s disease exposure risk in other settings [26,38,39]. Augmenting these data, the singular study analysed in the scoping review found that whilst multiple household members are involved in most livestock tasks, there is a clear pattern of exposure across age and gender lines.

Furthermore, Gizaw et al (2020) found that adult men dominate in slaughtering, marketing and herding, whilst adult women are more involved in feeding, cleaning barns, and managing sick animals. Young men primarily engage in marketing, assisting with breeding, and herding, while young women and children are more exposed through food hygiene and close contact with animals kept near or inside the home. These findings reflect both gender and age-specific responsibilities. Children’s close proximity to the home, especially girls, was thought to put them more at risk from diseases relating to chickens, animals kept at the household, and sanitation issues, whilst boys were considered to be more at risk of diseases relating to herding animals and staying away from the household.

Behavioural practices were also found to influence exposure risk. For instance, washing hands before eating was less frequent among young women, whilst handling contaminated tools was primarily done by adult men for non-zoonotic disease, and adult women for the transmission of zoonotic disease. These hygiene factors are crucial in the transmission of zoonotic and neglected tropical diseases [40–42].

Consumption habits further compound differential exposure. All household members were involved in disease transmission through the consumption of raw meat and milk, but proportional piling scores indicate that men had the most frequent or risky practices. Raw meat and milk consumption is often associated with the risk for zoonotic disease transmission [43,44] and is found to be associated with gender in Ethiopia in some studies [44,45]. The KIs noted that cultural norms and taboos, such as men being served first, consuming preferred cuts, or women not being allowed to eat certain foods, likely contribute to the gendered differences in consumption patterns.

#### Loss of nutrition and food security

Loss of nutrition, potentially leading to malnutrition, may result from reduced productivity and mortality of livestock, resulting in a decreased availability of animal-source foods (direct impact) or income loss, which reduces the capacity to purchase food (indirect impact). Although the study identified during our scoping review did not disaggregate nutritional impacts by gender, key informants and other studies suggest this impact is unlikely to be equal [45]. Augmenting KIs’ perception that women are the household member most impacted by nutritional loss, some KIs described a hierarchy at mealtimes, with men eating first and women only eating once the men were satisfied, as well as men eating better cuts of meat. The KIs also believed that the loss of nutrition in children was due in part to this feeding hierarchy, as well as the potential decrease in milk production from diseased livestock.

Women often rear livestock as a source of food and income for their families, rather than as a larger economic activity. Thus, loss of livestock owned and controlled by women could have a much greater impact on household nutrition and children in particular [18]. Children’s consumption of animal source foods, such as milk, is a critical source of protein and other essential nutrients, and their loss has been linked to stunting and undernutrition [46,47]. Whilst evidence is mixed as to whether boys or girls are more at risk from nutritional shocks [45,48,49], it is generally supported that women and girls are more vulnerable to malnutrition due to the prioritisation of men’s nutrition, the feeding hierarchy, and the loss of empowerment that women can face due to reduced incomes, hampering their access to resources [50].

### Psychological and emotional well-being

Previous research has shown that the psychological and social impacts of animal disease are frequently overlooked. Instances of animal disease can harm farmers’ mental health, leading to feelings of failure, grief, and financial distress, particularly when livestock death disrupts food security, household income, and perceived productivity [51–53]. The study identified by the scoping review collected data on the impacts of various livestock diseases and found that up to 6.4% of groups reported psychological/social impacts as a way these diseases impact households in Ethiopia. These findings align with results from KIIs which found that women particularly experienced psychological impacts. Among KIs, women were thought to face heightened emotional strain due to increased labour burdens, loss of decision-making power, and diminished status within the household following livestock disease events. These are stressors that can compound and amplify one another, reinforcing a cycle of disempowerment.

Prolonged exposure to stress and anxiety from such instances can result in very real indirect health impacts [54]. For instance, one study in Ghana found that 60% of livestock farmers exhibited poor mental health, with 72% showing signs of depression, 66% anxiety, and 59% stress as a direct result of livestock losses [52]. Livestock are more than just animals to many pastoralists; they are members of the family, and much loved [55]. Mental health decline can also result from knock-on pressures beyond grief, or social or economic concerns. Women may be at risk of developing poor mental health as a result of difficulties relating to increased labour burden and responsibilities, resource scarcity, and added pressures in sustaining their households and livestock (*ibid*).

Livestock are more than just tools for earning income and gaining nutrition. They also provide social benefits to farmers, through a show of wealth and status, use in cultural and religious ceremonies, and as offerings to guests [5]. Animal disease and the consequential impact on herds can also have a psychological impact, resulting from a reduction in status, shame, judgment from the community, and the toll this can take on people’s feelings of self-worth and community standing. Whilst the KIs only raised this as an impact for women in Ethiopia, psychological impacts can affect all household members. However, women’s mental health in agriculture is an area of growing research interest in the West, with increasing evidence that agricultural labour stress, coupled with household and invisible care expectations, results in increased instances of poor mental health among farming women compared to men [56,57].

### Livelihood impacts of animal disease

#### Increased and/or altered labour burden

Women in particular have the dual burden of unpaid reproductive work in caring for the home and family, in tasks such as preparing food, collecting water, feeding young children, breastfeeding, collecting cooking fuel, and cleaning, as well as paid and agricultural work. Increased labour burden was the impact most frequently raised by the KIs as affecting women, with particular reference made to the increased labour that results from the death of a donkey due to disease, and women’s role in caring for sick household members.

Donkeys play a vital role in women’s livelihoods, particularly in the collection of water and wood fuel from long distances [58]. Without the assistance of donkeys, KIs noted how women must do these tasks themselves. Women’s care of unwell household members limits their ability to spend time on income-generating activities and other requirements, and further overburdens women. Women have been found to spend between 11.85-14 hours per day working in Ethiopia, not inclusive of the increased labour burden that comes as a result of shocks such as livestock disease [27], leaving little time for additional activities.

The KIs also raised an increased labour burden, particularly affecting young girls, due to them often mirroring their mother’s role in the household, such as cleaning, cooking, and caring for small ruminants if adult women become unable to do so. Children have been found to spend 8 hours per day herding, watering animals, and collecting dung [27], which again suggests that many children do not have significant time for additional tasks, especially during the school term.

#### Reduction in women’s empowerment

Livestock production offers a unique opportunity to increase women’s empowerment by providing them with the ability to have ownership of assets, and a source of income they can control independently, while increasing their decision-making power. Increasing women’s livestock ownership can work to reduce the gender asset gap, with women who have greater assets found to increase spending on children’s education and health [59]. The literature particularly considers women’s empowerment in relation to its role in child nutrition, with women who are empowered in livestock production (or other income-generating activities) in possession of key resources and power to decide on household food purchases and consumption [60].

Conversely, women’s excessive labour burden has been found to be a leading cause of their disempowerment, another key burden faced by women as a result of livestock disease, limiting their ability to do other things, such as earn income or attend group meetings [61]. Findings from the KIIs largely align with the existing literature in this area, with KIs broadly agreeing that animal disease imposes a significant burden on women.

#### School absenteeism

Livestock disease can lead to school absenteeism due to reduced household income (particularly that of women), increased child labour requirements, and health impacts. The KIs noted these first two points, highlighting that livestock disease may increase labour requirements, resulting in an overall increase in labour burdens, while decreasing disposable income to pay for school. Complementing these findings, Gizaw et al. (2020) found that children dropping out of school was not a highly reported impact of disease by FDG participants; however, 25% reported it as an impact of anthrax, and 18.3% as an impact of foot-and-mouth disease, specifically.

Loss of income may lead to school absenteeism in two additional ways: it may mean parents are unable to pay school registration or exam fees, buy uniforms or other school needs, resulting in children being barred from attending classes until arrears are settled or fees can be met [5]; or that children are required to assist with alternative income sources to make up for reduced household income.

## Limitations

While this study presents a crucial first step in understanding the intrahousehold burden of animal disease in Ethiopia, several limitations should be acknowledged. Firstly, the scoping review revealed the extent of the lack of data in the literature that directly addresses the intrahousehold, gender-disaggregated impacts of animal disease in Ethiopia. Only one study met the inclusion criteria for a detailed intrahousehold analysis, significantly limiting the breadth and diversity of findings that could be drawn. Finally, while the key informants provided rich contextual insights, their expert opinions may reflect institutional bias, and the purposive sampling may limit the representativeness of the perspectives that were gathered.

## Conclusions

This study provides the first focused examination of the intra-household burden of animal disease in Ethiopia, combining expert insights from key informant interviews with a scoping review. We found a scarcity of evidence within the literature, with only one eligible study. Other studies exist that analyse divisions of labour or the roles of livestock, but only the included study directly links these to animal disease.

The limited literature and expert testimonies reveal that disease exposure and impacts are shaped by deeply embedded gendered divisions of labour, decision-making power, and access to resources. Women, men, girls and boys are not only exposed to disease to varying degrees through different livestock production and consumption practices, but also bear distinct economic, livelihood, and health and well-being consequences.

Our findings underscore that without gender and age-disaggregated data, the substantial but uneven burden of animal disease within households remains obscure, risking interventions that inadvertently reinforce inequalities. There is a significant need for further research that focuses on strengthening our understanding of aspects such as income control from livestock, household members’ resource allocation, decision-making power, benefit sharing, and livestock production practices, with a focus on gender and intrahousehold dynamics. Furthermore, this future research must move beyond household-level generalisations to capture the lived realities of individual members, integrating quantitative economic measures with qualitative social science approaches to uncover both direct and indirect impacts of animal disease fully.

Strengthening primary data collection systems that incorporate gender-sensitive design will enable programmes to develop targeted and contextually appropriate interventions. Such gender-specific interventions would enable more effective mitigation of human health risks, while reducing mortality and morbidity in livestock. In doing so, they would support and improve food security while protecting livelihoods, particularly for household members who are currently least visible in animal health burden data, yet most vulnerable to its consequences.

## Declarations

All authors declare that they have no conflicts of interest

## Data Availability

The search strings were adapted using keywords relating to intra-household, gender, disease, impact, livestock and Ethiopia for use in each database, available in the OSF Registries at https://doi.org/10.17605/OSF.IO/K5JEC.

